# A Symbolic Regression Approach to Hepatocellular Carcinoma Diagnosis Using Hypermethylated CpG Islands in Circulating Cell-Free DNA

**DOI:** 10.1101/2022.01.25.22269799

**Authors:** Rushank Goyal

## Abstract

**Purpose:** Hepatocellular carcinoma is the most common primary liver cancer, accounting for 90% of cases, and a major cause of death worldwide. Despite this, alpha-fetoprotein tests are the only blood-based diagnostic tools available, and their use is limited by their low sensitivity. DNA methylation changes, which have been implicated in a majority of cancers, offer an alternative method of diagnosis through measuring such changes in circulating cell-free DNA present in blood plasma.

**Method:** A genetic programming-based symbolic regression approach was applied to gain the benefits of machine learning while avoiding the opacity drawbacks of “black box” models. The data included plasma samples from 36 patients with hepatocellular carcinoma as well as a control group of 55 that contained patients with and without cirrhosis. A 75-25 train-test splitting was done before training.

**Results:** The symbolic regression methodology developed an equation utilizing the methylation levels of three biomarkers, with an accuracy of 91.3%, a sensitivity of 100%, and a specificity of 87.5% on the test data. The performance matches prior research while providing the added benefits of transparency.

**Conclusion:** Circulating cell-free DNA presents opportunities for minimally invasive early diagnosis of hepatocellular carcinoma, and utilizing transparent machine learning approaches like symbolic regression can allow accurate diagnosis by combining biological and mathematical principles. Future validation of the model obtained here on a larger and more diverse dataset can reveal the potential for such approaches in cancer diagnosis and pave the way for further research.

## 1 Introduction

Hepatocellular carcinoma (HCC) is the most common primary liver cancer, with approximately 782,000 new cases and 746,000 deaths yearly (Ferlay et al., 2014). It is the second largest cause of cancer deaths in East Asia and sub-Saharan Africa as well as being the fastest rising cause of cancer-related death in the United States (Rawla et al., 2018). Risk factors such as chronic Hepatitis B (CHB), chronic Hepatitis C (CHC), nonalcoholic steatohepatitis (NASH), and aflatoxin exposure contribute to the development of hepatocelluar carcinoma, with CHB and CHC alone accounting for 75% of cases (Chen, 2018; Baecker et al., 2018; Ahmed et al., 2019; Wang & Gribskov, 2019). With alpha-fetoprotein (AFP) tests being the only blood-based diagnostic tool currently available for HCC – the utility of which is hampered by its low sensitivity – there is an unmet need for an effective plasma test to enable early diagnosis (Xu et al., 2017).

CG dinucleotides, also known as CpG pairs, are under-represented in the human genome (21% of expected) due to the formation of methylcytosine through attachment of methyl groups to cytosine in CpG pairs; the methylcytosine spontaneously deaminates to thymine (Illingworth & Bird, 2009). There are, however, interspersions of largely nonmethylated sequences called CpG islands (CGIs) that possess large numbers of GC and CG nucleotides (Deaton & Bird, 2011). Hypomethylation or hypermethylation of these CGIs acts as a prevalent molecular signature for most cancers, including HCC (Wen et al., 2015). In a number of studies, such methylation changes were detected years before other signs of cancerous development (Shi et al., 2007).

Circulating cell-free DNA (cfDNA) is a term describing extracellular DNA found in body fluids like blood, sputum, urine, etc. (Sun et al., 2019). Its importance and promising outlook as a non-invasive marker for cancer has been recognized, utilizing genetic, methylation, and, to a lesser extent, quantitative analyses (Aarthy et al., 2015; Kustanovich et al., 2019). For HCC in particular, methylation analysis of cfDNA has yielded multiple diagnostic biomarkers, including but not limited to p15, p16, GSTP1 and RASSF1A (Ng et al., 2018).

Symbolic regression is a machine learning (ML) technique that attempts to create a mathematical expression to explain the target variable utilizing a range of basic mathematical functions, and, specifically, genetic programming-based symbolic regression (GPSR) is an implementation of SR that uses genetic programming to search the massive SR solution spaces effectively (Wilstrup & Kasak, 2021). GPSR has been able to perform well in deriving expressions for real-world applications in fields as disparate as biology, robotics, physics and finance (Orzechowski et al., 2018). Evidence suggests that it outperforms other ML techniques on smaller datasets, which is useful in clinical and biological contexts where data is often limited (Wilstrup & Kasak, 2021). Its transparency compared to other “black box” machine learning models also makes it more suited to application in clinical settings (Quinn et al., 2021; Price, 2018).

## 2 Materials and Methods

The data used for the analysis were obtained from the NCBI Gene Expression Omnibus with the accession number GSE63775. The data consist of plasma samples from 55 control subjects, out of which 17 had cirrhosis, and 36 HCC patients (Wen et al., 2015). Values for each CGI in the samples are measured in methylated alleles per million mapped reads (MePM). Stratified random train-test splitting was performed in a 75-25 ratio (random seed of 1) to obtain the training and testing sets respectively.

Python 3.7.12 was used, with pandas and numpy for preprocessing, matplotlib and seaborn for plotting and visualization, scikit-learn for train-test splitting and model evaluation, and gplearn for training the symbolic classifier model. A symbolic classifier works by first developing a symbolic regressor and then passing the output through a logistic function to produce a prediction value, which corresponds to 0 (negative) if less than 0.5, and 1 (positive) otherwise. The symbolic classifier was trained using population_size=2000 and parsimony_coefficient=0.01, with a random seed of 1 to ensure reproducibility (Andrew L. Beam, 2020).

## 3 Results

Figure 1 shows the output expression in a tree form. Equation 1 shows the final model as a standard mathematical equation, where *chr*_5_, *chr*_21_ and *chr*_19_ represent the levels of the three CGIs identified as biomarkers by GPSR – chr5:92923487-92924497, chr21:40757602-40757900 and chr19:41531804-41532051, respectively. *x* is the output, and can be put through the logistic function to obtain the probability (*p*) of hepatocellular carcinoma, as shown in Equation 2. A probability less than 0.5 indicates that the absence of HCC is more likely, and vice versa.

**Figure 1:**
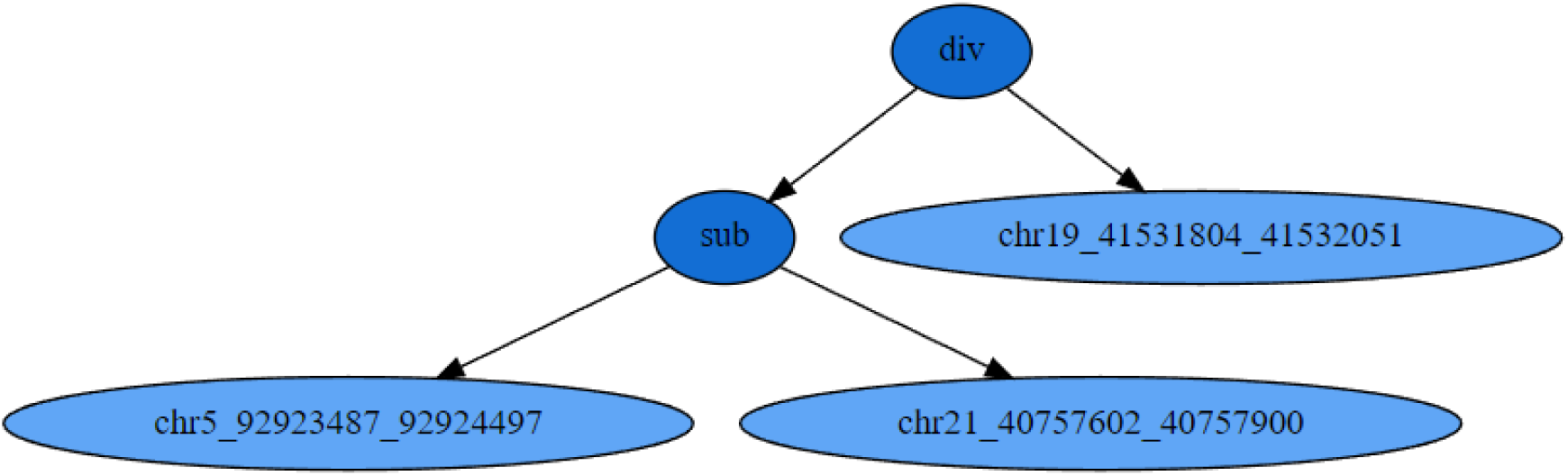
Output Equation as Tree

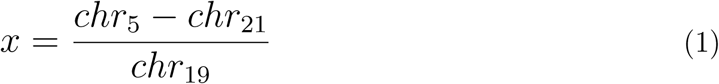

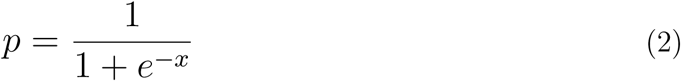

The results when utilizing this equation for classification on the testing dataset are shown in Figure 2 in the form of a confusion matrix. The accuracy comes out to be 91.3%, and the sensitivity and specificity are 100% and 87.5%, respectively.

**Figure 2:**
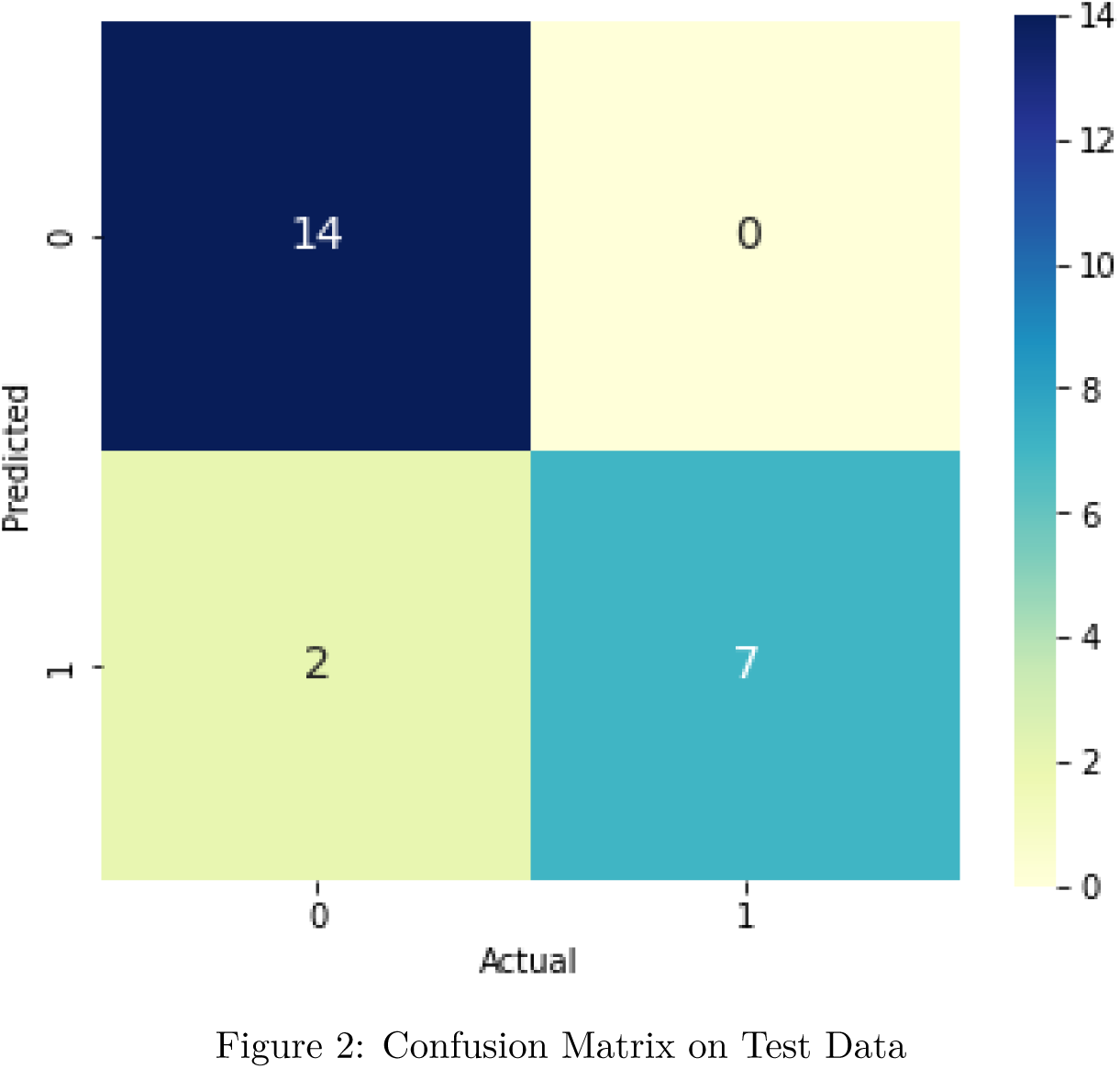
Confusion Matrix on Test Data

## 4 Discussion

Prior ML work in this area has attained similar performances, though it lacks the explainability of symbolic regression (Filho et al., 2020; Khalid et al., 2015). Utilizing a combination of random forest, the least absolute shrinkage and selection operator (LASSO) and logistic regression, Xu et al. (2017) obtained a sensitivity of 83.3% and a specificity of 90.5% on their validation set. Zhang et al. (2020) were able to get a sensitivity and specificity of 91.93% and 100% respectively using 11 biomarkers identified by a support vector machine (SVM) model. Another study also used an SVM to achieve a precision (positive predictive value) of 96% and a recall (sensitivity) of 86% (Gonçalves et al., 2021).

Multiple studies have identified blood plasma biomarkers without applying ML. Lin et al. (2005) showed that around 77% of HCC patients possess p16 methylation and 41% possess DAPK methylation. 55.7% of patients have a methylated CCND2 gene (Tsutsui et al., 2010). RASSF1A hypermethylation was present in 42.5% of patients, correlating with tumor size (Yeo et al., 2005). Ji et al. (2004) identified MT1M and MT1G as biomarkers with high specificities of 93.5% and 87.1% respectively, though their sensitivities were low, at 48.8% and 70.2% respectively.

Compared to simple biomarker analysis, machine learning techniques display better performance due to their more complex nature. The symbolic regression approach described in this paper, however, attains similar sensitivity and specificity to conventional ML approaches, is much more transparent and allows for mathematical reasoning of the results in a biological context, which can be a source of useful further research (Narayanan et al., 2022; Cardoso et al., 2020).

Since the data in this study contained cfDNA – which can be obtained from blood plasma – rather than analyzing liver tissue, the results present possibilities for minimally invasive diagnosis of HCC. Indeed, cfDNA holds potential for improved early cancer diagnosis in general (Stewart et al., 2018). Further analysis of the results on a larger and more diverse validation set as well as biological analyses of the three-biomarker signature are important, but the results obtained here show initial promise.

## Data Availability

All data utilized are available online through the NCBI GEO under the accession number GSE63775.

https://www.ncbi.nlm.nih.gov/geo/query/acc.cgi?acc=GSE63775

## 5 Data Availability Statement

The data are available in the National Center for Biotechnology Information’s Gene Expression Omnibus (NCBI GEO) under the accession number GSE63775.

## 7 Ethical Declarations

This research study was conducted retrospectively using human subject data made available by prior research. Ethical approval was not required, as confirmed by the institutional review board (IRB) of the Sri Aurobindo Institute of Medical Sciences in Indore, India.

